# A Predictive Autoantibody Signature in Multiple Sclerosis

**DOI:** 10.1101/2023.05.01.23288943

**Authors:** Colin R. Zamecnik, Gavin M. Sowa, Ahmed Abdelhak, Ravi Dandekar, Rebecca D. Bair, Kristen J. Wade, Christopher M. Bartley, Asritha Tubati, Refujia Gomez, Camille Fouassier, Chloe Gerungan, Jessica Alexander, Anne E. Wapniarski, Rita P. Loudermilk, Erica L. Eggers, Kelsey C. Zorn, Kirtana Ananth, Nora Jabassini, Sabrina A. Mann, Nicholas R. Ragan, Adam Santaniello, Roland G. Henry, Sergio E. Baranzini, Scott S. Zamvil, Riley M. Bove, Chu-Yueh Guo, Jeffrey M. Gelfand, Richard Cuneo, H.-Christian von Büdingen, Jorge R. Oksenberg, Bruce AC Cree, Jill A. Hollenbach, Ari J. Green, Stephen L. Hauser, Mitchell T. Wallin, Joseph L. DeRisi, Michael R. Wilson

**Affiliations:** UCSF Weill Institute for Neurosciences, Department of Neurology, University of California, San Francisco, CA, USA; Department of Medicine, McGaw Medical Center of Northwestern University, Chicago, IL, USA; UCSF Weill Institute for Neurosciences, Department of Psychiatry and Behavioral Sciences, University of California, San Francisco, CA, USA; Department of Biochemistry and Biophysics, University of California, San Francisco, CA, USA; Department of Epidemiology and Biostatistics, University of California, San Francisco, CA USA; Veterans Affairs, Multiple Sclerosis Center of Excellence, Washington, DC and University of Maryland School of Medicine, Baltimore, MD, USA; Chan Zuckerberg Biohub, San Francisco, CA, USA

**Keywords:** Multiple sclerosis, antigen, phage display, PhIP-Seq, serology, neurofilament light chain

## Abstract

Although B cells are implicated in multiple sclerosis (MS) pathophysiology, a predictive or diagnostic autoantibody remains elusive. Here, the Department of Defense Serum Repository (DoDSR), a cohort of over 10 million individuals, was used to generate whole-proteome autoantibody profiles of hundreds of patients with MS (PwMS) years before and subsequently after MS onset. This analysis defines a unique cluster of PwMS that share an autoantibody signature against a common motif that has similarity with many human pathogens. These patients exhibit antibody reactivity years before developing MS symptoms and have higher levels of serum neurofilament light (sNfL) compared to other PwMS. Furthermore, this profile is preserved over time, providing molecular evidence for an immunologically active prodromal period years before clinical onset. This autoantibody reactivity was validated in samples from a separate incident MS cohort in both cerebrospinal fluid (CSF) and serum, where it is highly specific for patients eventually diagnosed with MS. This signature is a starting point for further immunological characterization of this MS patient subset and may be clinically useful as an antigen-specific biomarker for high-risk patients with clinically- or radiologically-isolated neuroinflammatory syndromes.

## Introduction

Multiple sclerosis (MS) is a chronic inflammatory autoimmune disease that primarily affects the white matter of the central nervous system.^1,2^ While traditionally thought to be T-cell mediated,^3,4^ the widespread success of B cell depleting therapies in humans has focused attention on a central role of B cells in the etiology and progression of MS.^5,6^ MS is often disabling when untreated; it affects women more often than men, and appears to have increased in frequency, with nearly 1 million currently affected in the US alone.^7,8^

In the large majority of patients, MS presents as a bout or relapse: a neurological symptom complex suggestive of demyelination such as optic neuritis, partial myelitis, brainstem syndrome or multifocal onset.^1,2,4^ MRI of the CNS performed shortly after clinical onset often shows acute demyelinating plaques that enhance following intravenous contrast administration.^9^ In addition, there are often one or more chronic demyelinating lesions,^9^ suggesting that neuroinflammation precedes symptomatic onset.^4^ The recognition of the “radiologically isolated syndrome” (RIS) demonstrated that plaques could be identified on brain and spinal cord MRI prior to symptom onset when these scans were performed for other reasons.^10^ Histopathologically, demyelinating plaques are heterogeneous and are populated by complex and varying immune cell populations, including CD4^+^ and CD8^+^ T cells, activated macrophages, B cells and deposition of immunoglobulin and complement.^3,4,11^

Several retrospective studies that assess clinical records found that subclinical neurologic episodes occur more frequently in persons who received an MS diagnosis later in life, suggesting that MS prodrome involves an ongoing inflammatory process.^12–15^ Given that MS lesions are thought to develop after demyelinating events, it may be that these subclinical neurological episodes are indicative of ongoing neuroinflammatory process in the presymptomatic period.^16^ Although molecular evidence for this has been limited, the theory that neuronal tissue injury precedes clinical onset is supported by recent studies showing elevated serum neurofilament light (sNfL), indicating axonal damage several years before MS diagnosis.^17–19^

Studies in a number of autoimmune diseases – systemic lupus erythematosus,^20^ type 1 diabetes mellitus,^21^ and rheumatoid arthritis,^22^ among others - indicate that diagnostic autoantibodies can appear years before the onset of symptoms.^23^ In MS, by contrast, no such validated diagnostic autoantibodies exist.^24^ Indeed, the role of autoantibodies in MS,^5,25^ and their link to pathogenesis,^6,26^ has proven notoriously difficult to ascertain, with studies suggesting myelin or potassium channel (KIR4.1) as putative antigenic targets.^6,25,27,28^ A hallmark of MS is the presence of unique oligoclonal bands in the cerebrospinal fluid of almost all PwMS,^16,26,29^ which implies intrathecal antibody synthesis.^30^ However, no clearly predictive or diagnostic autoantigen has been identified.^24^ The need for prospectively ascertained cohorts of sufficient size to study this heterogeneous disease makes prospective autoantigen identification exceptionally challenging.^31,32^

To assess autoantibody signatures, we used a large prospective incident MS cohort assembled during the Gulf War era (GWEMSC) from over 10 million individuals in the active-duty United States military population.^33^ From this group of cases, the Department of Defense Serum Repository (DoDSR) staff retrieved the earliest serum aliquot from entry into active duty, an average of 5 years before their first clinical symptom (**Fig. 1a,b**) and then another serum sample on average 1 year after this first attack. Control samples, without an MS diagnosis, were selected and matched to cases on age, sex, race/ethnicity, and dates of serum collection (year). After an unbiased autoantibody screen and sNfL measurements on these samples, (**Fig. 1c,d**), autoantibody results were orthogonally validated on both serum and cerebrospinal fluid (CSF) using a gold-standard prospective, incident MS cohort at the University of California, San Francisco (ORIGINS) that enrolled patients at clinical onset.

**Figure 1.**
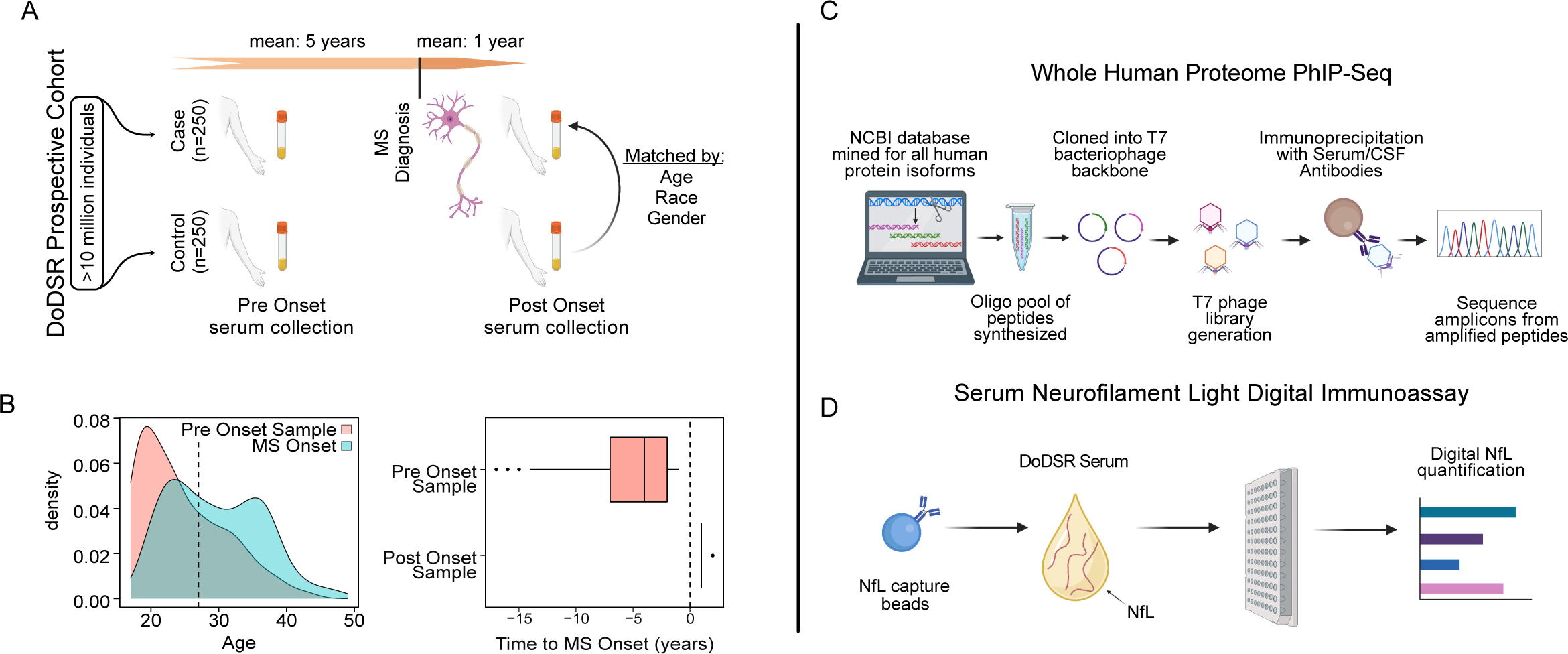
Overview of MS Biomarker study. A) Schematic of DoDSR cohort & collection. B) Age and time to symptom onset for MS cases. C,D) Molecular biomarker assays performed on DoDSR cohort of longitudinal sera.

## Results

### DoDSR and ORIGINS Multiple Sclerosis Cohorts

The DoDSR cohort contained 250 MS patients 5-years (5.0 ± **SD 3.3**) before and 1-year (1.2 ± **SD 0.4**) after first symptom onset and 250 non-MS healthy controls who were matched for age, sex, race/ethnicity, and serum collection dates (Table 1). The UCSF ORIGINS cohort, comprised of untreated patients presenting with their first-ever attack, consisted of 103 patients who were ultimately diagnosed with RRMS (Table 2). Other neurologic disease (OND) controls included 14 patients who enrolled in ORIGINS but were ultimately found to have a non-MS diagnosis (varicella zoster virus meningitis n=1, neuromyelitis optica spectrum disorder (NMOSD) n=4, vitamin B12 deficiency n=2, Leber’s hereditary optic neuropathy n=2, neurosarcoidosis n=2, primary CNS vasculitis n=1, lymphoma n=1) as well as 9 additional OND controls (migraine n=4, brainstem stroke n=1, anti-phospholipid antibody syndrome n=1, brainstem demyelinating syndrome n=1, idiopathic spastic paraparesis n=1, peripheral neuropathy n=1).

### DoDSR longitudinal serum shows distinct immunologic signature years before first relapse

Molecular profiling of both autoantibody repertoire and neuronal damage were carried out on the longitudinal samples acquired from the DoDSR (**Fig. 2**). sNfL, a marker of axonal injury, was measured in cases and controls at both timepoints. Among the pre-symptomatic serum samples, NfL levels were higher at time points closer to date of diagnosis (**Fig. 2b**). Consistent with a recent study,^17^ sNfL levels were significantly higher in post-onset samples compared to their pre-onset counterparts for PwMS (**Fig. 2c**).

**Figure 2.**
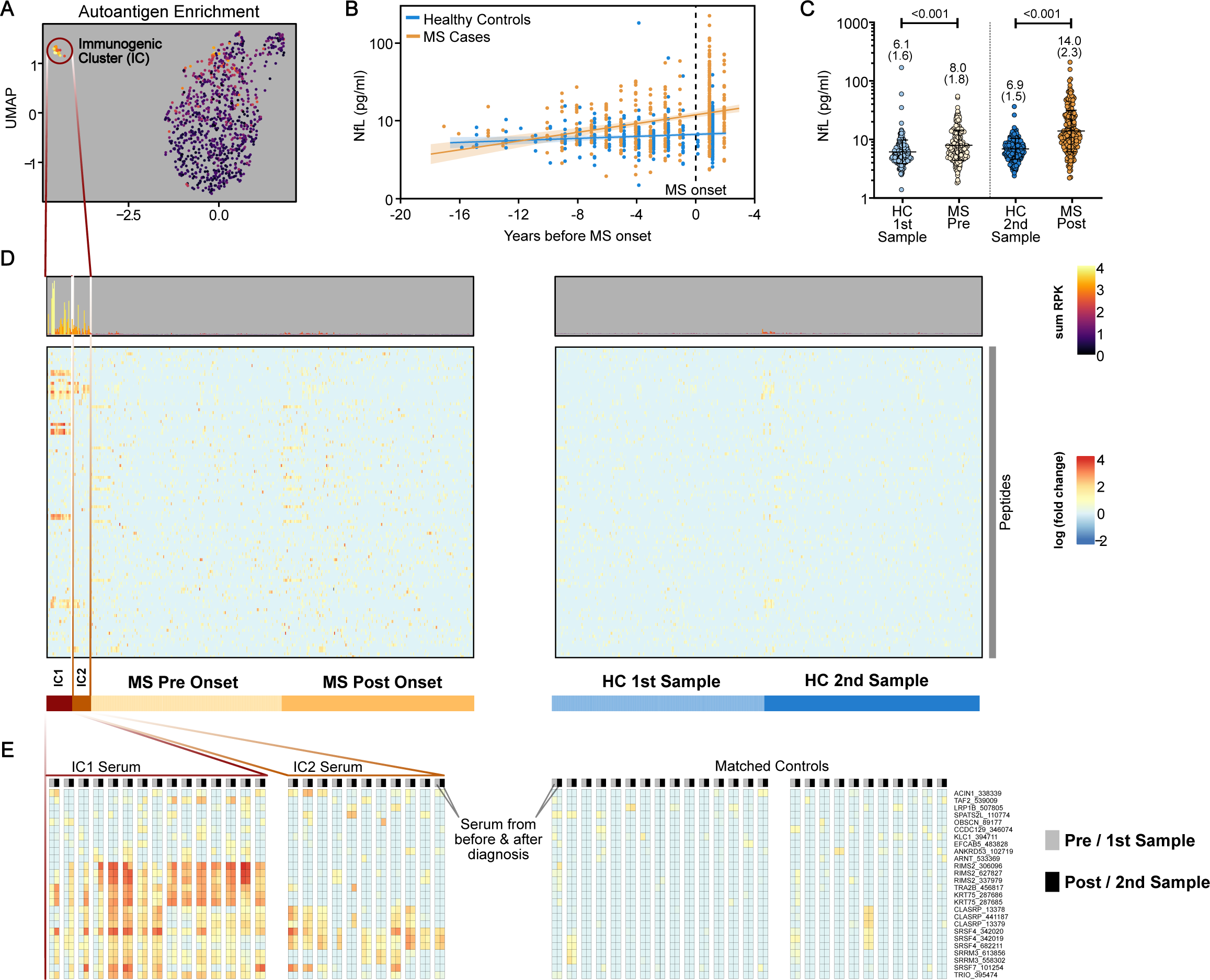
Profiling DoDSR MS Cohort. **A)** UMAP of autoantigen enrichments in PhIP-Seq screen of DoDSR sera, showing distinct immunogenic clusters (IC1 and IC2). **B)** sNfL levels across DoDSR cohort sera with respect to time before onset and **C)** grouped by time point for cases and controls. **D)** Enrichments of top 192 peptides, with IC patients pre and post onset samples highlighted on left grouped by hierarchical clustering. **E)** Blowout of top 26 most enriched peptides in IC patients grouped by gene, and respective enrichments in their non-MS matched controls.

In patients ultimately diagnosed with MS, sNfL levels were higher even many years before their first clinical flare when compared to the matched control cohort (**Fig. 2c**). Significant differences in sNfL levels across time points were not observed in the non-MS control cohort. Together, these data support evidence that at least some MS patients exhibit early signs of neuroaxonal injury long before onset of symptoms.

A whole-human proteome seroreactivity approach (PhIP-Seq)^34^ was employed to determine if an antigen-specific signature of future disease accompanied elevated sNfL levels. This technique uses a T7 phage display library to probe antibody-antigen interactions by immunoprecipitation from patient serum or CSF and has been used for unbiased biomarker detection in diseases of unknown etiology.^34–41^

The longitudinal nature of this cohort with its well-matched non-MS control samples allows for detection of changes in the antigenic repertoire before and after clinical onset. The collection of peptides enriched by immunoprecipitation using sera from patients (the autoantibody signature) in the cohort was consistent over time (**Supporting Fig. 1)**. When blinded, time points from the same patient could be unambiguously associated with nearly 96% accuracy across the 500 subjects assayed (**Supporting Fig. 2**). This consistency over time was present regardless of diagnosis. No compelling enrichment of antigens specific for PwMS post-diagnosis were observed.

We then asked whether there might be a serologic signature in patients that go on to develop MS compared to those who did not. To visualize the differences between the overall PhIP-seq results across all samples, a Uniform Manifold Approximation and Projection (UMAP) was generated using the fold-change enrichment for each peptide, relative to the control cohort (see Methods). In this projection, a distinct immunogenic cluster (IC) emerged that included PwMS before and after disease onset (**Fig. 2a**).

After unsupervised clustering of this cohort of samples, serum antibodies from this group of patients enriched several of the same peptides that were preferentially found in patients who developed MS (n=27) as opposed to controls without MS (n=3). These cases separated into two clusters: IC1, with a more polyspecific profile; and IC2 that appeared to be reactive to a subset of IC1 peptides. (**Fig. 2d**). These patients were grouped into an MS_IC_ category for the purpose of further analysis compared to those that did not exhibit reactivity (MS_no-IC_).

Patients in these clusters were defined by a clear but disparate group of enriched peptides derived from 54 different proteins in both pre- and post-onset samples (see Methods). Most striking about this result was its longitudinal stability (**Fig. 2e**). Analysis of all IC samples, both before and after disease onset, clearly shows that this class of antigens is enriched in both time points in the majority (17/26, 65%) of cases (one case did not meet sequencing depth cut-off in respective pre-symptomatic sample).

Alignment of peptides preferentially enriched by sera from the IC subset of individuals revealed a characteristic protein motif (“IC motif”) described by the regular expression *P-(SA)-x-(SGA)-R-(SN)-(LRKH)*, with the initial proline being the most conserved (**Fig 3a**). This motif is broadly represented in the human proteome (**Fig. 3a**). While the motif-bearing peptides derive from diverse coding sequences, proteins containing splicing activation and mRNA binding domains, due to their characteristic arginine-serine repeat sequences,^42–44^ i.e. SRSF4, SRRM3, and CLASRP were highly represented in the enriched set.

**Figure 3.**
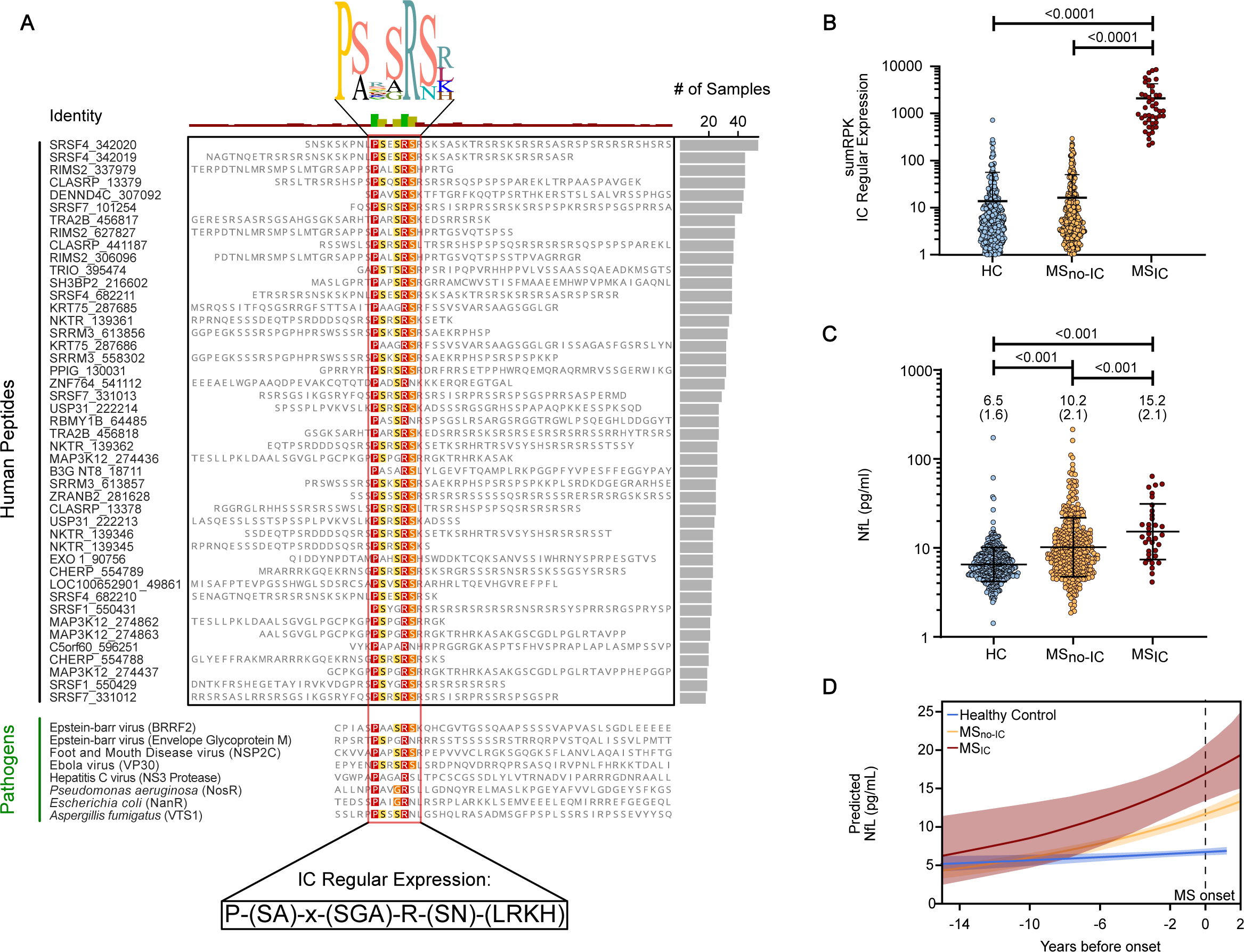
IC Peptide and Cohort Analysis. **A)** Protein alignment of peptides within human proteome library that contain the IC regular expression, in order of patient prevalence that exceeded cutoffs in either pre or post onset samples. Bottom is alignment of regular expression to selected pathogens that infect humans (via Prosite Scan). **B)** Normalized reads assigned to regex-containing peptides in MS patients within either IC cluster as compared to those without and controls. **C)** Serum levels of NfL in MS patients within IC clusters as compared to those without and controls. **D)** Serum NfL levels modeled over time for each group with respect to time to onset.

Searching the PROSITE database^45^ using the IC motif regular expression and not simply a most-probable collapsed sequence, disclosed the motif’s presence across many phyla. In addition to humans, the IC motif is present in several human pathogens, including Epstein-Barr virus (EBV) (both in the BRRF2 tegument protein and the envelope glycoprotein M), hepatitis C virus (NS3 protease), *Pseudomonas aeruginosa* (NosR), *Escherichia coli* (HTH-type transcriptional repressor) and *Aspergillus fumigatus* (RNA-binding protein vts1) (**Fig. 3a**).

Neither patient demographic nor clinical features (e.g., time to symptom onset, MS subtype and gender) distinguished these IC reactive patients from the larger cohort except for a possible association with race (p<0.164) (Table 3). Nevertheless, patients with enriched signal across peptides described by the IC motif had significantly higher sNfL levels than MS patients not assigned to this cluster, as well as the non-MS controls (**Fig. 3b-d)**.

### IC signature present in both cerebrospinal fluid and serum of patient subset at time of symptom onset in the independent, prospective ORIGINS cohort

While the longitudinal stability of this signature is important in the context of surveillance, we sought to replicate this finding in patients with incident MS or clinically isolated syndrome (high risk for MS) within 30 days of their first symptomatic episode, where its diagnostic value would be more apparent. Here, 126 paired CSF and serum samples were analyzed from a completely independent incident MS cohort of treatment-naïve patients (UCSF ORIGINS Study) of which most (n=103, 88%) were eventually diagnosed with MS after radiologic and clinical follow-up. A similar fraction of confirmed MS patients (8/103) expressed the same signature as those of IC reactive patients in the DoDSR longitudinal cohort, with characteristic enrichment of peptides containing the IC motif (**Fig 4a, b**). These patients similarly split into IC1 and IC2 type profiles, clustered together via UMAP (**Fig. 4d**) and demonstrated enrichment of these peptides in both compartments (**Fig. 4c**). Interestingly, this signature was highly specific for MS cases, where only one patient diagnosed with NMOSD having low-level enrichment in their serum but not in CSF, out of 23 confirmed OND controls. This result was orthogonally validated using a bead-based (Luminex) multiplexed indirect immunofluorescence assay featuring four distinct peptides that were highly enriched across the MS_IC_ patient samples (**Fig. 5a**). Relevant peptides from proteins RIMS2, KRT75, SRSF4, and TRIO were included, each of which contained variations of the IC motif. After testing all ORIGINS paired CSF and sera using this assay, only patients exceeding the cut-off were ultimately diagnosed with MS (**Fig. 5b-e**).

**Figure 4.**
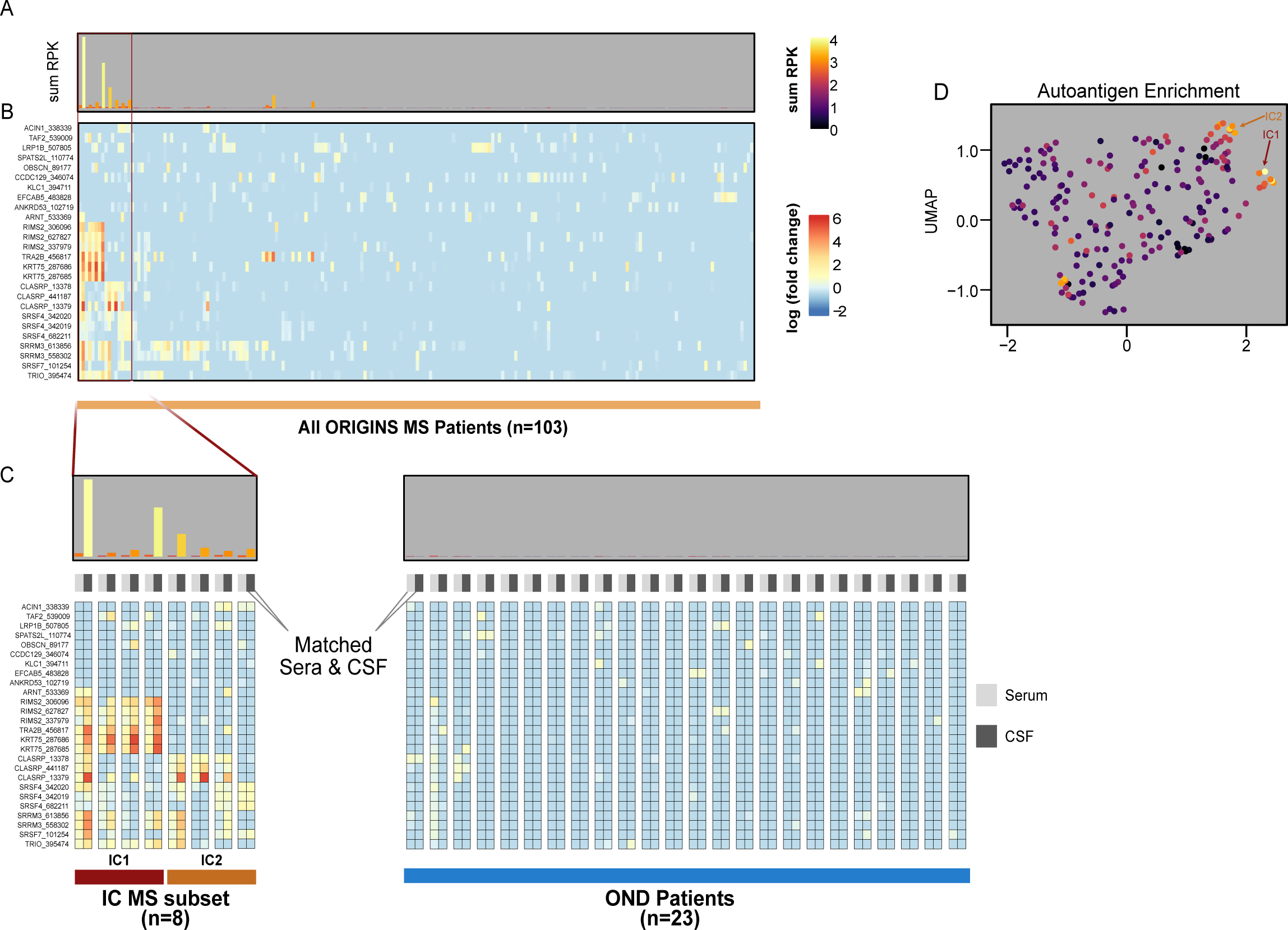
PhIP-Seq Characterization of ORIGINS validation cohort in CSF and sera. **A)** sum of normalized reads for previously identified top 26 peptides and **B)** PhIP-Seq enrichments of ORIGINS patient sera and CSF for peptides mapping to same regions of ICs from DoDSR cohort, relative to controls. **C)** Blowup of IC signature patients with all OND controls with CSF and serum for each patient grouped by column. **D)** UMAP of PhIP-Seq enrichments for all ORIGINS patients, shaded by sum RPK levels from (A) Arrows indicate where IC clusters lie on UMAP.

**Figure 5.**
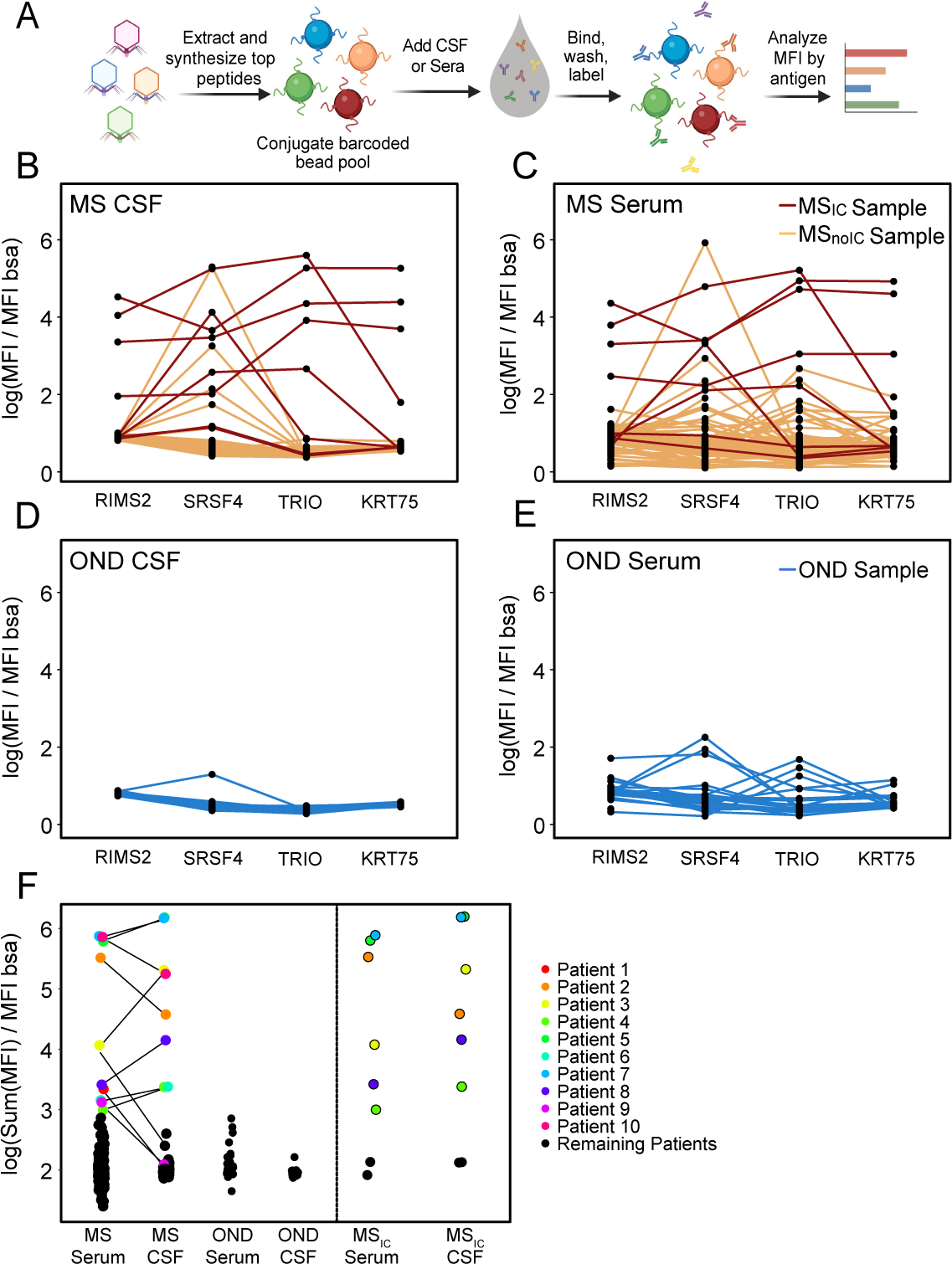
Luminex validation assay of selected IC peptides. **A)** Schematic for barcoded immunofluorescence assay against representative peptides from IC motif. Normalized median fluorescence intensity for each patient, matched by colored line, across all ORIGINS patients **B)** CSF and **C)** sera (n=103), as well as **D), E)** OND controls (n=23). **F)** Sum of MFI across peptides shown for all four groups, with those exceeding cutoff (normalized logMFI >3) in either CSF or sera highlighted and matched by patient (left). Patients identified by PhIP-Seq IC (right) highlighted separately.

Similar to the PhIP-Seq results, some degree of heterogeneity was observed across the peptide combinations, but samples from the same group of patients yielded enrichment of the IC motif (**Fig. 5b-e, Supporting Fig. 3**). A metric of the summed mean fluorescence intensity (MFI) (**Fig. 5f**) across these peptides confirmed and extended the PhIP-seq findings. MS_IC_ patients exhibited 6.7- and 8.2-fold increased normalized MFI compared to MS_no-IC_ in sera and CSF, respectively. Specifically, seroreactivity to peptides bearing the IC motif were highly specific for MS patients at the time of symptom onset, were not found in OND controls, and were nearly always present in both CSF and serum in each case.

To evaluate whether HLA variation may contribute to the unique phenotype observed in MS_IC_ patients, the classical HLA genes HLA-A, HLA-B, HLA-C, HLA-DPB1, HLA-DRB1, HLA-DQB1 were fully sequenced and genotyped (Supplemental Table 1). Due to limited sample size, formal analysis to determine whether any HLA locus or allele exhibited a significant association with MS_IC_ status was not possible. Qualitative assessment of allele frequencies shows that some HLA alleles were found at higher frequencies in the MS_IC_ group, relative to the remaining ORIGINS MS cohort; however, it must be noted that many alleles in the MS_IC_ were present as only single copies (Supplemental Table 2). Interestingly, all but one of the MS_IC_ individuals carry the HLA-DPB1*04:01 allele (Supplemental Table 1). This high carrier frequency of 83%, relative to the 67% observed in the remaining cohort, suggests a possible enrichment for the HLA-DPB1*04:01 allele in the MS_IC_ subset. However, these observations must be considered strictly speculative, as a much larger sample will be required to determine whether any observed differences in HLA frequency distributions between these groups is significant.

## Discussion

In this study, an MS-specific autoantibody signature was identified in a subset of patients years before their first clinical MS attack. This same signature was validated using two different methods in an independent, prospective cohort of patient samples (serum and CSF) collected near the time of their first MS relapse (UCSF ORIGINS cohort).

This study presents some of the first prospective autoantigen-specific biomarkers found in the prodromal phase of MS, which is consistent with prior clinical analyses that MS pathophysiology may begin years before symptom onset and diagnosis. The evidence includes numerous studies that thoroughly investigated clinical and indirect markers of this phase via chart review of health care claims.^14,31,46^ Documented symptoms range from neuropsychiatric phenomena such as fatigue and headache, to mental health dysfunction including anxiety and depression, and prescriptions for antispasmodics, all of which have elevated odds ratios in patients that went on to develop MS. Evidence from a similar prospective military cohort found that poor cognitive performance was found at a higher rate in patients who went on to receive an MS diagnosis in the following two years compared to controls.^47^

This work validates and adds to prior evidence of neuro-axonal injury occurring in patients during the MS prodromal phase,^17^ as sNfL is a robust biomarker that is likely specific to neuro-axonal damage and is elevated in the serum of MS patients compared to healthy controls.^19,48–50^ Elevated levels are associated with faster progression to EDSS scores of >4, and reduction in sNfL concentration is associated with initiation of disease-modifying therapy.^51^ In this study, significantly elevated sNfL levels in the pre-onset group of patients compared to matched controls were observed, especially in the distinct immunogenic cluster we identified, supporting the notion that neurodegeneration is already occurring as a result of underlying immune-mediated neuroaxonal pathology in the prodromal phase of MS.

The role of autoantibodies in MS remains unclear; many candidate autoantigens have not survived validation studies.^24^ However, other rare demyelinating diseases of the CNS such as NMOSD or myelin oligodendrocyte glycoprotein antibody-associated disease (MOGAD) were originally part of the MS spectrum, and were definitively separated only after the identification of disease-specific autoantibodies,^52^ findings that also informed subsequent therapeutic development.^53,54^

Remarkably, prior studies screening smaller numbers of MS patient serum or CSF samples on human and viral antigen libraries uncovered a similar motif as described here in MS_IC_ patients.^39,55^ The identification, corroboration, and orthogonal confirmation of this autoantibody signature in two distinct, large patient cohorts across three different disease epochs (i.e., presymptomatic, initial flare and post-diagnosis), strongly suggests that the MS_IC_ signature has clinical diagnostic potential, especially in the context of early MS detection. Given its robust specificity for MS both before and after diagnosis, an autoantibody serology test against the MS_IC_ peptides could be implemented in a surveillance setting for patients with high probability of developing MS, or crucially at a first clinically isolated neurologic episode.

The similarity of this motif to domains contained in a broad array of infectious agents, including two EBV proteins,^39,55^ suggests it is possible that infection by one or several of these agents contributes to an autoreactive response and disease pathogenesis, perhaps via molecular mimicry.^56,57^ The role of infection as the temporally initiating event in MS etiology is the focus of current work^18,58^ and is crucial to the differentiation of causal versus spuriously associated features of the MS prodrome. However, this remains challenging given near-ubiquitous seropositivity for many of the implicated viruses, including EBV, while only a small fraction of those infected develop MS. Nonetheless, EBV infection, and infectious mononucleosis in particular, represents the most compelling epidemiologic link to MS,^59–61^ including from the DoDSR studied here.^18^ Given the degree of polyspecificity of this motif in the human proteome, it will be important to elucidate the precise origins of the MS_IC_ signature with special attention to past exposures, genetic risk factors, and temporal dynamics of disease manifestation.

This study has several limitations. The small number of patients identified in this cluster makes genetic or other associations difficult. While similarity exists with this motif and those found in other human infectious agents such as EBV, this study did not directly measure titers of antiviral antibodies. The validation panel used in the Luminex assay consists of only four peptides that encompass the motif, and thus sensitivity would likely be improved by adding more candidates to the multiplexed assay.

## Conclusion

This work identifies a longitudinally stable autoantibody profile that is present before, during, and after the time of first symptom onset for a subset of MS patients. Taken together with elevated sNfL levels, this hallmark patient cluster is an attractive target for further immunological and clinicopathologic study. This study, along with other evidence of ongoing neurodegeneration during MS prodrome, suggests additional evaluations of these patients beyond sNfL and autoantibodies might bear additional insight into underlying immunological processes during this crucial disease phase and stratify patients with different immunological features.

## Supporting Information

Supporting information contains sample fingerprinting, pre- and post-onset PhIP-Seq similarity data for MS cohort and matched controls, and data on RPK vs MFI agreement with respective peptides for ORIGINS cohort patients.

## Data Availability

All data produced in the present study are available online at

https://github.com/UCSF-Wilson-Lab/MS_DoD_and_ORIGINS_study_data

## Acknowledgements

The authors would like to acknowledge the Chan Zuckerberg Biohub and their sequencing team for sequencing assistance. The authors would also like to thank the patients and their families for their contribution to this study.

## Funding and Disclosures

This work was supported by the Valhalla Foundation (SLH, MRW, JRO, BAC), the Westridge Foundation (MRW, HCvB, AJG), NINDS R35NS122073 (SLH, MRW and RD), National Multiple Sclerosis Society RFA-2104-37504 (MRW, MTW, JH, BAC, CRZ), NMSS RFA-2104-3747 (JRO), Bruce Sachs, the UCSF Dean’s Office Medical Student Research Program (GMS), Chan Zuckerberg Biohub (JLD, SAM), John A. Watson Scholar Program, UCSF (CMB), Hanna H. Gray Fellowship, Howard Hughes Medical Institute (CMB), NIH NIH R01AI158861 (JAH and KJW) and the University of California President’s Postdoctoral Fellowship Program (CMB).

MRW receives unrelated research grant funding from Roche/Genentech and Novartis, and received speaking honoraria from Genentech, Takeda, WebMD and Novartis.

## Author Contributions

CRZ, GMS, MRW, MTW and JLD designed the experiments. CRZ performed all PhIP-Seq and Luminex assays; RDB performed the Luminex assays, and GMS and SM performed PhIP-Seq immunoprecipitations and NGS library preparation. SAM, CF and AEW assisted with DoDSR sample preparation. AA, NJ and KA performed the sNfL studies, and AA analyzed the data. RD and CMB built the analysis code and pipeline for PhIP-Seq data, and analyzed the PhIP-Seq data with GMS, as well as generating figures. KCZ, AT, CF, JA, CG, EBT, HCvB, RPL, RG and ELE curated and collected patient samples and clinical data for the UCSF ORIGINS cohort. KJW, JRO and JAH performed the HLA sequencing, genotyping and association. MTW generated the DoDSR cohort and epidemiologic analysis, and AJG contributed valuable sNfL analysis interpretation. SJH, BACC, JRO, together with the UCSF MS-EPIC Team and JMG, CYG and RC, led the recruitment and phenotyping of the UCSF ORIGINS cohort. SJH, BACC and JLD gave valuable experimental guidance and manuscript editing. All authors contributed to study design. CRZ and MRW wrote the manuscript. All authors provided editorial comments.

## Patient Cohort Details

### DoDSR Cohort

The incident Gulf War Era MS cohort (GWEMSC, n=2,691) was used as the source for identifying US military MS cases in this study.^62^ The GWEMSC cohort was drawn from the broader US military population that served during the Gulf War era (1990-2007) with relevant demographic and clinical data abstracted from Department of Defense (DoD) and Department of Veterans Affairs (VA) records.^63^ This cohort is population-based within the US military and thereby has a male preponderance and race and ethnicity subgroups that mirror the US census. All veterans in this cohort are service-connected for MS, which requires evidence of clinical signs upon examination attributable to MS during or within seven years after active duty military service. Additionally, all cases were adjudicated by the VA MS Center of Excellence study team and met the McDonald MS criteria.^64^

The DoD Serum Repository (DoDSR) MS cohort (IRB-1624644-4) was created using the clinical information on cases from the GWEMSC and linking serum aliquots before and after first symptom onset from the DoDSR. To build the DoDSR MS cohort, a stratified population-based sample was created within the GWEMSC. Based on prespecified demographic strata, the DoDSR staff identified a group of MS cases (n=250) that had at least one serum aliquot in the repository prior to MS onset and one aliquot after. The earliest serum sample obtained before onset of MS symptoms from each Veteran with MS was identified by DoDSR staff along with a second sample within two years after initial MS symptoms. For each MS case, one healthy control (n=250) was randomly selected and matched on year of entry to the military, age, sex, race, ethnicity, and dates of blood collection (within 180 days).

### ORIGINS Cohort

All RRMS and clinically isolated syndrome (CIS) subjects were participants in either the University of California, San Francisco (UCSF, IRB 14-15278) ORIGINS study and were diagnosed according to the 2017 McDonald criteria.^64^ Subjects were not on immunomodulatory or immunosuppressive disease-modifying therapy at the time of sample collection.

Additional CSF experimental reference controls for ORIGINS PhIP-Seq analysis were generated from 22 healthy or non-MS participants enrolled in a biobanking study “Immunological Studies of Neurologic Subjects”. Additional serum experimental reference controls for ORIGINS PhIP-Seq analysis were generated from 95 reference donors provided by the New York Blood Center.

## Methods

### Nfl Measurements

NfL values were measured in duplicate from sera samples with sufficient volumes (n= 944) using the Simoa® NF-light™ Advantage Kit (Quanterix®, Massachusetts, United States) on an HD-X analyzer by lab assistants blinded to the clinical characteristics of the included subjects. Samples with a coefficient of variation (%CV) >20% or where duplicate values were not available were excluded from the final analysis (n= 14). The mean intra-run %CV of the included samples were (n=930, 5.02% (±3.9)), while the inter-run %CV for the quality control samples were 9.42% and 9.02% for low and high concentrations, respectively.

### PhIP-Seq Protocol

Phage Immunoprecipitation Sequencing was performed as previously described with some modifications for a high throughput protocol.^65^ 96-well, 2mL deep well polypropylene plates were incubated with a blocking buffer (3% BSA in TBST) overnight at 4°C to prevent nonspecific binding. Blocking buffer was then replaced with 500 µL of freshly grown phage library (at 10^11^ pfu/mL) and 1 µL of human sera or 10 µL of CSF in 1:1 storage buffer (PBS supplemented with 0.04% NaN_3_, 40% Glycerol, 40mM HEPES). To facilitate antibody-phage binding, the deep well plates with library and sample were incubated overnight at 4°C on a rocker platform in secondary containment. 20 µL of each of Pierce Protein A and G Beads (ThermoFisher Scientific, 10002D & 10004D) slurry were aliquoted per reaction and washed 3 times in TNP-40 (140mM NaCl, 10mM Tris-HCL, 0.1% NP40). After the final wash, beads were resuspended in TNP-40 in half the slurry volume (20uL) and added to the phage-patient antibody mixture and incubated on the rocker at 4°C for 1 hour. Beads were then transferred to a vacuum manifold compatible filter plate^66^ then washed five times with RIPA (140mM NaCl, 10mM Tris-HCl, 1% Triton X, 0.1% SDS) using an Integra Viaflow96 to add RIPA each time and using the vacuum to remove the supernatant. In each wash, beads were incubated for 5 minutes with gentle rocking at 4°C. After the fifth wash, immunoprecipitated solution was resuspended in 150 µL of LB-Carb and then added to 0.5mL of log-phase BL5403 *E. coli* for amplification (OD_600_ = 0.4-0.6) until lysis was complete (approximately 2h) on a 800 rpm shaker. After amplification, sterile 5M NaCl was added to lysed E. coli to a final concentration of 0.5M NaCl to ensure complete lysis. The lysed solution was spun at 3220 rcf for 20 minutes and the top 500 µL was filtered through a 0.2 mM PVDF filter plates (Arctic White, #AWFP-F20080) to remove remaining cell debris. Filtered solution was transferred to a new pre-blocked deep-well plate where patient sera was added and subjected to another round of immunoprecipitation and amplification. The final lysate was spun at 3220 rcf for 30 minutes, with supernatant then filtered and stored at 4°C for subsequent NGS library prep.

Phage DNA from each sample was barcoded and amplified (Phusion PCR, 18 rounds) and subjected to Next-Generation Sequencing on an Illumina NovaSeq Instrument (Illumina, San Diego, CA) at an average read depth of 1 million reads per sample. Two post onset MS samples and one second time point control sample library did not pass sequencing quality control and were excluded from analysis.

### Luminex Assay

Peptides containing the relevant motif from TRIO, KRT75, RIMS2 and SRSF4 (Table 4) were synthesized by LifeTein LLC and conjugated to Bovine Serum Albumin (BSA) via SMCC coupling to cysteines on BSA at a 1:1 final mass ratio. Spectrally distinct Luminex beads were conjugated in separate 1.5mL protein lo-bind tubes to their corresponding BSA-peptide antigen. A control bead population was made by conjugating BSA only (Sigma-Aldrich, cat# 10735094001). Each bead conjugation was performed at a concentration of 5 μg of protein per 1 million beads in a 0.5mL reaction volume. Conjugation was done via an EDC/sulfo–N-hydroxysuccinimide coupling strategy to free amines using Ab coupling kit following manufacturer’s instructions (Luminex, catalog 40-50016), as performed previously.^65^

**Table 4.** Peptides used in Luminex assay. Red refers to motif segment that maps to regular expression.

| Protein Name | Amino Acid Sequence |
| --- | --- |
| Keratin 75 (KRT75) | SRRGFSTTSAIT<u>PAAGRSRFS</u>SVSVARSAAGS |
| Regulating Synaptic Membrane Exocytosis 2 (RIMS2) | PSLMTGRSAPPS<u>PALSRSHPR</u>TGSVQTSPSST |
| Serine/arginine-rich splicing factor 4 (SRSF4) | SRSRSNSKSKPNL<u>PSESRSRS</u>KSASKTRSRSKSR |
| Trio Rho guanine nucleotide exchange factor (TRIO) | GA<u>PSTSRSR</u>PSRIPQPVRHHPP |

All serological analyses were performed in duplicate, and beads were pooled on the day of use. Thawed serum samples were diluted in PBS + 0.05% Tween 20 (PBST) containing 2% nonfat milk and mixed with pooled protein-coated beads (2000–2500 beads per protein) at a final serum dilution of 1:250, or final CSF dilution of 1:20. Samples were incubated for 1 hour at room temperature at 250rpm, washed thrice with PBST, then stained with 1:2000 phycoerythrin conjugated anti–human IgG Fc Ab (BioLegend, catalog 637310) in PBST for 30 minutes at room temperature. Beads were washed twice with PBST and analyzed in a 96-well plate format on a Luminex LX 200 cytometer. The net MFI for each peptide antigen was calculated for each sample by dividing by the MFI of that samples’ corresponding intra-assay BSA negative control and averaging across duplicate wells.

### Sequencing and HLA genotyping

For each sample, 100 ng of high quality DNA was fragmented with the Library Preparation Fragmentation Kit 2.0 (Twist Bioscience, San Francisco, USA). Following fragmentation, DNA was repaired and poly-A tails were attached and ligated to Illumina-compatible dual index adapters with unique barcodes. After ligating, fragments were purified with 0.8X ratio AMPure XP magnetic beads (Beckman Coulter, Brea, USA). Double-size selection was performed (0.42X and 0.15X ratios) and libraries of approximately 800 bp were selected, at which point libraries were amplified and purified using magnetic beads.

Following fluorometric quantification, each sample was pooled (30 ng/sample) via ultrasonic acoustic energy. The Twist Target Enrichment kit (Twist Bioscience, San Francisco, USA) was then used to perform target capture on pooled samples. Sample volumes were then reduced using magnetic beads and DNA libraries were bound to 1394 biotinylated probes. Probes were designed specifically to target all exons, introns and regulatory regions of the classical HLA loci, including HLA-A, HLA-B, HLA-C, HLA-DPB1, HLA-DRB1 and HLA-DQB1. Then, streptavidin magnetic beads were used to capture fragments targeted by the probes. Captured fragments were then amplified and purified. Bioanalyzer (Agilent, Santa Clara, USA) was then used to analyze the enriched libraries. After evaluation, enriched libraries were sequenced using a paired-end 150 bp sequencing protocol on the NovaSeq platform (Illumina, San Diego, USA). Following sequencing, HLA genotypes were predicted using HLA Explorer (Omixon, Budapest, Hungary).

### Statistics and Analysis

#### sNfL Analysis

Independent samples student t-test (equal variance not assumed) was applied to compare log-transformed NfL concentrations between samples collected from people with multiple sclerosis and age- and gender-matched healthy controls, at each timepoint separately. To compare between longitudinal NfL concentration in MS_IC_ and MS_NO-IC_ groups, mixed linear models assessed log-transformed NfL concentrations as dependent variables in the three groups (HC, MS_IC_, MS_NO-IC_) correcting for age at sampling (fixed effects). All reported p-values were Bonferroni-adjusted.

Predicted NfL concentrations used to plot their dynamics in the three groups over the duration between the first and second samples were fitted using a linear mixed model of NfL concentration (log-transformed) accounting for the group (HC, MS_IC_, MS_NO-IC_), duration between samples in relation to documented MS onset, with an additional interaction term (groups x duration). The statistical analysis was conducted with SPSS® (version 28) and JMP® Pro (version 16).

#### PhIP-Seq

Raw sequencing reads were aligned to a reference library using Bowtie2 as described previously.^65^ To correct for differences in sequencing depth, read counts for each peptide within a sample were normalized to the total reads expressed as reads per 100,000 (RPK). Fold-change for each peptide was generated from the mean RPK of the controls, and a z-score was calculated from the background distribution.

A custom bioinformatics pipeline was used to generate a list of candidate peptides enriched in the MS group. For each peptide, fold-change (FC) values for all MS samples were calculated by dividing the RPK value by the mean RPK of all matched healthy control samples. FC values were then used to calculate z-score values. The first round of filters identified enriched peptides with a minimum RPK >= 1 and a FC >= 10. Among enriched peptides, more stringent cut-offs were applied to identify candidate peptides. This included FC >= 100 and z-scores >= 10. Mutually enriched peptides from the same gene were also added to the candidate peptide list if they had FC >= 100 and a z-score >= 3. Additionally, peptides from given genes were only included in the candidate list if they had a sum rpK >= 50. A 7 amino acid (AA) k-mer analysis was performed on all enriched peptides. Enriched peptides that shared an identical 7AA sequence with at least one other peptide were kept for further analysis.

Since a bonafide autoantibody would likely target the same epitope on multiple enriched peptides, the candidate list was further narrowed by filtering out peptides without k-mer overlaps. Given the size of the cohort and the tendency of individuals to enrich highly personalized epitopes, we further narrowed the candidate list requiring peptides to have a fold-change > 10 in at least 15/250 MS cases.^36^

#### Defining patient clusters for DoD cohort

When we performed hierarchical clustering of the resulting candidate peptides, we found that a subset of MS pts and two healthy controls (n = 18) enriched several peptides bearing a nearly identical epitope. Unsupervised clustering with uniform manifold approximation and projection (UMAP) was used to further define patient clusters among enriched candidate peptides. Among samples in the DoD cohort, all MS and healthy control (HC) samples were clustered only using 192 enriched peptides from 151 proteins which were a product of the cutoffs stated above. Separately, a smaller panel of enriched peptides was used to further define sub-clusters. To assemble this panel of peptides, for each peptide both the sum rpK across all MS patients and sum rpK across all HC samples was calculated. These values were used to calculate a sum rpK ratio of MS to HC. Peptides with a sum rpK > 800 across all samples and an MS to HC ratio > 6 were kept in our smaller target panel of 13 peptides (9 proteins). For each sample, both the sum rpK of our target panel as well as the sum rpK for all 119 peptides bearing the characterized motif were used.

Unsupervised clustering with UMAP (n_neighbors = 20, n_components = 3, learning_rate = 1, init = “random”, n_epochs = 100000) revealed one outlier cluster of 49 samples which contained increased expression in both of our target peptide panels. This subset of samples was re-clustered under the same parameters, which created two patient clusters. Cluster 1 contained 30 samples (n = 18) which showed higher sum rpK values among our target panel relative to the 19 samples in cluster 2 (n = 12).

#### Defining patient clusters for ORIGINS cohort

The same custom bioinformatics pipeline was used to identify enriched peptides among CSF and serum samples within the ORIGINS cohort. To identify the top enriched peptides in the CSF and serum, the sum FC of samples with FC > 10 was calculated. In total, this filtered out 2,307 peptides in the CSF (sum FC > 10) and 289 peptides (sum FC > 12) in the serum. A panel of 26 peptides that were enriched in 9 or more DoD samples (Fig 1B) was used to identify patient clusters. The sum rpK of this panel was calculated across all samples. Hierarchical clustering was used to identify a subset of 16 samples (n = 8) with the highest sum rpK were filtered. This subset of samples was divided into two clusters due to the difference in the FC expression profile among the panel of 26 peptides. Cluster 1 (n = 4) showed more pronounced expression in RIMS2, TRA2B, KRT75, SRRM3 and TRIO. Cluster 2 (n = 4) showed more pronounced expression in CLASRP and SRSF4.

We used the following cut-offs in our analysis code (package, PairSeq) to generate the candidate list of peptides:

MIN_RPK = 1

ZSCORE_THRESH1 = 3

ZSCORE_THRESH2 = 10

FC_THRESH1 = 10

FC_THRESH2 = 100

SUM_RPK_THRESH = 50

## Notes

### Competing Interest Statement

MRW receives unrelated research grant funding from Roche/Genentech and Novartis, and has received speaking honoraria from Genentech, Takeda, WebMD and Novartis.

### Author Declarations

IRB of University of California, San Francisco gave ethical approval for this work (14-15278) IRB of Veterans Affairs Medical Center gave ethical approval for this work (IRB-1624644-4)

### Summary of Updates

Error in author name in citation 39.

