## Supplementary material for "A Predictive Autoantibody Signature in Multiple Sclerosis": Demographic Tables (from another version of this preprint: version 1, 2023-05-02)

**Table 1 - DoDSR cohort demographics.**

| | MS Cases | Healthy Controls | Overall |
|---|---|---|---|
| | (N=250) | (N=250) | (N=500) |
| Gender | | | |
| Female | 84 (33.6%) | 84 (33.6%) | 168 (33.6%) |
| Male | 166 (66.4%) | 166 (66.4%) | 332 (66.4%) |
| Race and Ethnicity | | | |
| White | 159 (63.6%) | 159 (63.6%) | 318 (63.6%) |
| Black | 86 (34.4%) | 86 (34.4%) | 172 (34.4%) |
| Hispanic Ethnicity | 5 (2.0%) | 5 (2.0%) | 10 (2.0%) |
| Birth year | | | |
| Mean (SD) | 1970 (8.0) | 1970 (8.0) | 1970 (8.0) |
| Time between samples (years) | | | |
| Mean (SD) | 6.18 (3.3) | 6.07 (3.4) | 6.12 (3.3) |
| MS Subtype | | | |
| Relapsing Remitting | 192 (76.8%) | 0 (0%) | 192 (38.4%) |
| Secondary Progressive | 43 (17.2%) | 0 (0%) | 43 (8.6%) |
| Primary Progressive | 14 (5.6%) | 0 (0%) | 14 (2.8%) |
| Progressive Relapsing | 1 (0.4%) | 0 (0%) | 1 (0.2%) |

| | | | |
|---|---|---|---|
| Post Onset Serum Sample Collection (years*) | | | |
| Mean (SD) | 1.2 (0.4) | NA | 1.2 (0.4) |
| *Time in years from first symptom onset to serum collection | | | |

**Table 2 - ORIGINS cohort demographics.**

| ORIGINS cohort | | |
|---|---|---|
| | MS (n=103) | OND (n=23) |
| Sex | | |
| Female, n (%) | 67 (65.0%) | 14 (60.9%) |
| Male, n (%) | 36 (35.0%) | 9 (39.1%) |
| Age, yrs (mean ± SD ) | 34.1 ± 7.4 | 39.9 ± 13.4 |
| Race | | |
| White, n (%) | 74 (71.8%) | 15 (65.2%) |
| Black, n (%) | 4 (3.9%) | 2 (8.7%) |
| Asian, n (%) | 9 (8.7%) | 1 (4.3%) |
| Ethnicity | | |
| Latino, n (%) | 8 (17.5%) | 3 (13.0%) |
| Not Latino, n (%) | 95 (82.5%) | 20 (87%) |
| EDSS (median [IQR] {Range}) | 2 [1.5, 2.9] {0-6.5) | |
| Disease duration, months (median [IQR] {Range}) | 1 [0, 1] {0-19} | |

**Table 3 - $MS_{IC}$ demographic analysis.**

| | $MS_{IC}$ | $MS_{no-IC}$ | P-value |
|---|---|---|---|
| | (N=27) | (N=223) | |
| Race and Ethnicity | | | |
| White | 21 (77.8%) | 138 (61.9%) | 0.164 |
| Black | 5 (18.5%) | 81 (36.3%) | |
| Hispanic Ethnicity | 1 (3.7%) | 4 (1.8%) | |
| Sex | | | |
| Female | 9 (33.3%) | 75 (33.6%) | 1.0 |
| Male | 18 (66.7%) | 148 (66.4%) | |
| Age at First Symptom Onset (years) | | | |
| Mean (SD) | 28.6 (7.0) | 29.7 (6.8) | 0.5 |
| Median [Min, Max] | 29.0 [18.0, 42.0] | 29.0 [18.0, 49.0] | |
| MS Subtype | | | |
| Relapsing Remitting | 24 (88.9%) | 168 (75.3%) | 0.5 |
| Secondary Progressive | 2 (7.4%) | 41 (18.4%) | |
| Primary Progressive | 1 (3.7%) | 13 (5.8%) | |
| Progressive Relapsing | 0 (0%) | 1 (0.4%) | |
| Most Recent DSS score | | | |
| Mean (SD) | 3.11 (1.7) | 3.57 (2.3) | 0.2 |
| Median [Min, Max] | 3.0 [0, 7.0] | 3.00 [0, 9.0] | |
| Post Onset Serum Sample Collection (years*) | | | |

| | | | |
|---|---|---|---|
| Mean (SD) | 1.3 (0.5) | 1.2 (0.4) | 0.5 |
| Median [Min, Max] | 1.0 [1.0, 2.0] | 1.00 [1.0, 2.0] | |
| *Time in years from first symptom onset to serum collection | | | |
