## Supporting Data Figures for "A Predictive Autoantibody Signature in Multiple Sclerosis" (from another version of this preprint: version 1, 2023-05-02)

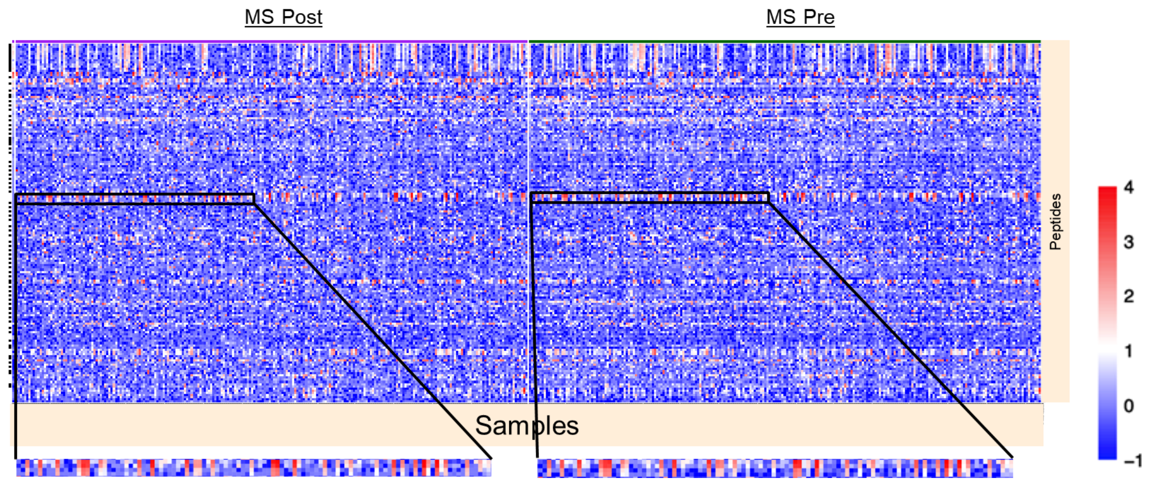


Supporting Figure 1. Heatmap of logRPK values from whole proteome PhIP-Seq data on MS patient sera from DoDSR cohort. Top peptides shown with peptide enrichment blowout to demonstrate similarity between pre and post onset sample enrichments over time.

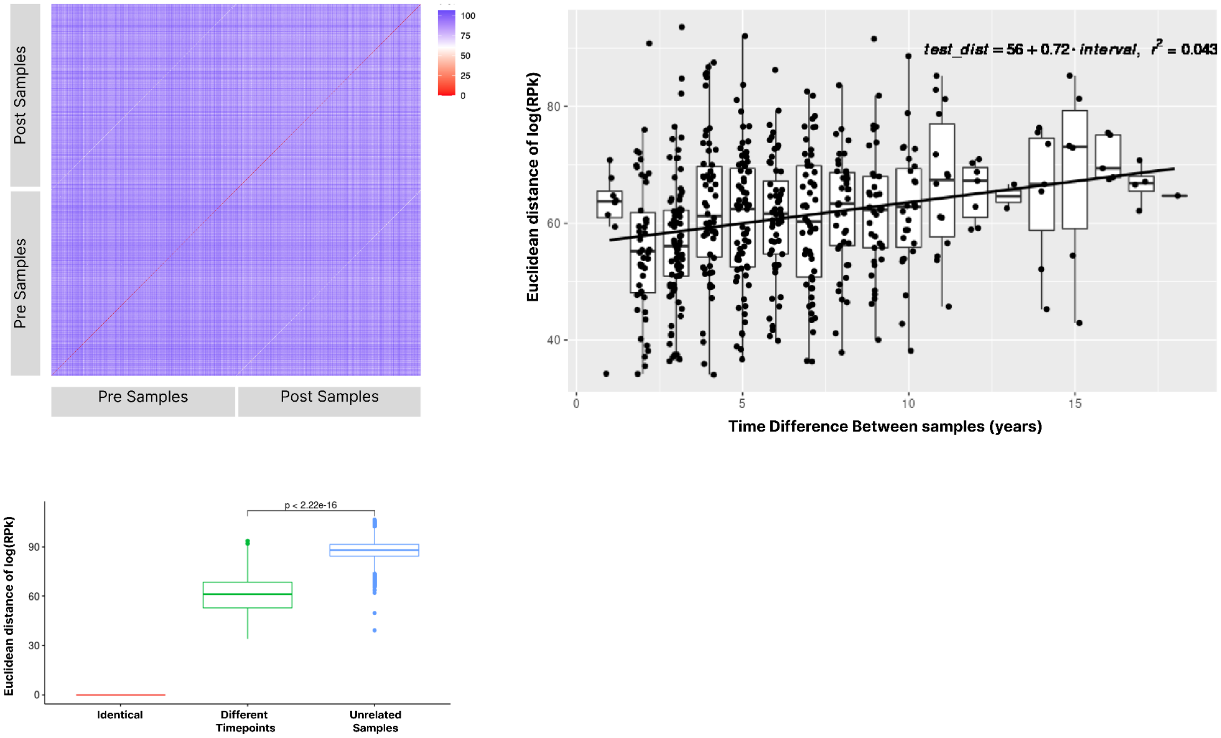


Supporting Figure 2. (A) Unfiltered normalized read counts (rpK) were log transformed, and a correlation matrix was computed using the Euclidean distance measure. The similarity between any two samples was defined by their n-dimensional Euclidean distance, where each peptide in the library was treated as a dimension. The rows and columns of this matrix are ordered such that the early timepoints are followed by the late timepoints. The central diagonal stripe in the heatmap represents a distance of zero where the distance was computed between the same sample. However, the diagonal stripes represent distances calculated between different timepoints of the same sample. (B) Box plots of the Euclidean distance between timepoints, with a trend towards increasing distance at longer intervals. However, patterns of self-reactivity remain stable for more than 15 years. (C) A boxplot demonstrates the significant difference in Euclidean distance between two timepoints of the same sample versus the distance between two unrelated samples.

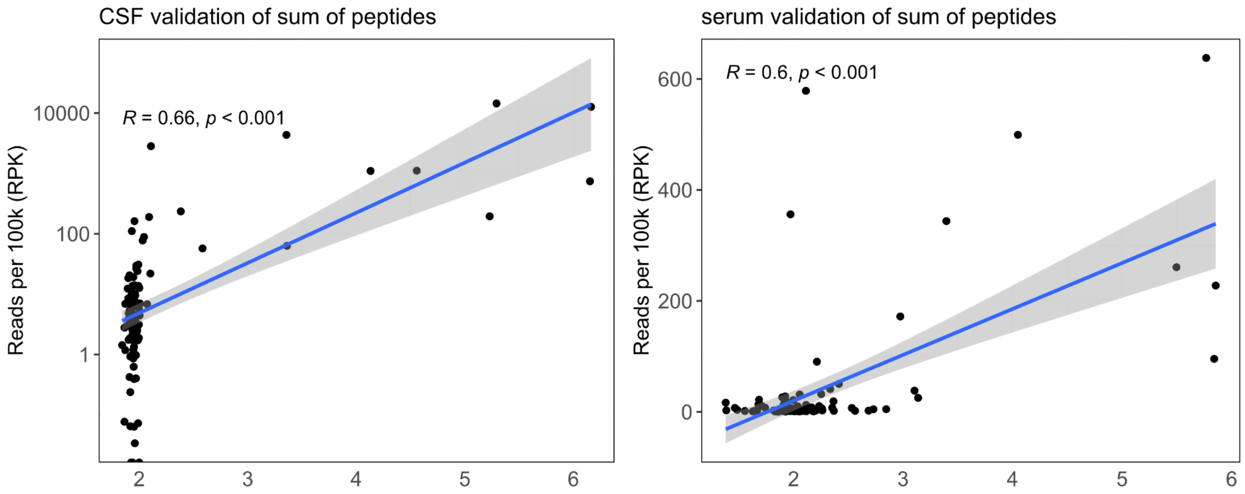


Supporting Figure 3. Correlation of sum of MFI from all four peptides of interest in Table 4 from Luminex assay with their sum of respective normalized read counts from PhIP-Seq data in both CSF and serum samples from ORIGINS cohort.
